# T cell assays differentiate clinical and subclinical SARS-CoV-2 infections from cross-reactive antiviral responses

**DOI:** 10.1101/2020.09.28.20202929

**Authors:** Ane Ogbe, Barbara Kronsteiner, Donal T. Skelly, Matthew Pace, Anthony Brown, Emily Adland, Kareena Adair, Hossain Delowar Akhter, Mohammad Ali, Serat-E Ali, Adrienn Angyal, M. Azim Ansari, Carolina V Arancibia-Cárcamo, Helen Brown, Senthil Chinnakannan, Christopher Conlon, Catherine de Lara, Thushan de Silva, Christina Dold, Tao Dong, Timothy Donnison, David Eyre, Amy Flaxman, Helen Fletcher, Joshua Gardner, James T. Grist, Carl-Philipp Hackstein, Kanoot Jaruthamsophon, Katie Jeffrey, Teresa Lambe, Lian Lee, Wenqin Li, Nicholas Lim, Philippa C. Matthews, Alexander J. Mentzer, Shona C. Moore, Dean J Naisbitt, Monday Ogese, Graham Ogg, Peter Openshaw, Munir Pirmohamed, Andrew J. Pollard, Narayan Ramamurthy, Patpong Rongkard, Sarah Rowland-Jones, Oliver Sampson, Gavin Screaton, Alessandro Sette, Lizzie Stafford, Craig Thompson, Paul J Thomson, Ryan Thwaites, Vinicius Vieira, Daniela Weiskopf, Panagiota Zacharopoulou, Oxford Immunology Network Covid-19 Response T cell Consortium, Oxford Protective T cell Immunology for COVID-19 (OPTIC) Clinical team, Lance Turtle, Paul Klenerman, Philip Goulder, John Frater, Eleanor Barnes, Susanna Dunachie

## Abstract

A major issue in identification of protective T cell responses against SARS-CoV-2 lies in distinguishing people infected with SARS-CoV-2 from those with cross-reactive immunity generated by exposure to other coronaviruses. We characterised SARS-CoV-2 T cell immune responses in 168 PCR-confirmed SARS-CoV-2 infected subjects and 118 seronegative subjects without known SARS-CoV-2 exposure using a range of T cell assays that differentially capture immune cell function. Strong *ex vivo* ELISpot and proliferation responses to multiple antigens (including M, NP and ORF3) were found in those who had been infected by SARS-CoV-2 but were rare in pre-pandemic and unexposed seronegative subjects. However, seronegative doctors with high occupational exposure and recent COVID-19 compatible illness showed patterns of T cell responses characteristic of infection, indicating that these readouts are highly sensitive. By contrast, over 90% of convalescent or unexposed people showed proliferation and cellular lactate responses to spike subunits S1/S2, indicating pre-existing cross-reactive T cell populations. The detection of T cell responses to SARS-CoV-2 is therefore critically dependent on the choice of assay and antigen. Memory responses to specific non-spike proteins provides a method to distinguish recent infection from pre-existing immunity in exposed populations.

## Introduction

In late 2019 the new virus severe acute respiratory syndrome coronavirus 2 (SARS-CoV-2) emerged, causing the range of clinical diseases known as COVID-19^1, 2^. While the majority of SARS-CoV-2 infections are asymptomatic or result in mild disease, some individuals develop severe respiratory symptoms which may result in hospital admission and death leading to high global mortality^3, 4^, especially older adults and those with comorbidities^5^. Understanding the immune responses resulting from exposure to SARS-CoV-2, and distinguishing these from the responses made to seasonal coronaviruses, is a pre-requisite to defining immune correlates of infection and protection against subsequent SARS-CoV-2 disease. This in turn is centrally important in comparing with protective vaccine-induced immunity and may contribute to future public health policies including shielding advice.

Antibody responses to SARS-CoV-2 are important but remain complex. In a recent large-scale study of healthcare workers, PCR-confirmed SARS-CoV-2 infection resulted in measurable antibodies after 20 days in nearly all participants, with high specificity^6^. However, there is wide variability. Other studies have reported that antibodies may be absent early in disease, levels of neutralising antibodies are highly variable^7^, and antibody titres wane over time^8^. In contrast, studies of SARS-CoV infection indicate that T-cell responses may be more durable^9^. A number of studies have demonstrated the presence of T cell responses to the virus during acute disease and in recovery. Using *in silico* predicted HLA-class I and II peptide pools, CD4+ T cell responses to SARS-CoV-2 were demonstrated in all subjects who had recovered from COVID-19 and CD8+ responses were demonstrated in 70%^10^. This study also found T cell reactivity to SARS-CoV-2 epitopes in 50% of archived samples from pre-pandemic (2015-2018) subjects using a 24-hour activation induced markers (AIM) assay. Additionally, a Swedish study demonstrated a highly activated cytotoxic phenotype in acute disease and vigorous polyfunctional T cell responses in convalescent subjects^11^. Interestingly, the latter study reported T cell responses to SARS-CoV-2 in seronegative household contacts, which may represent either seronegative infection or pre-existing cross-reactive immune memory to seasonal coronaviruses.

The role of prior exposure to human seasonal coronaviruses including alpha coronaviruses (HCoV-NL63 and HCoV-229E), and beta coronaviruses (HCoV-HKU1 and HCoV-OC43), that may generate SARS-CoV-2 cross-reactive T cell immune responses, is of substantial interest. Whilst prior exposure to the original SARS-CoV and to MERS-CoV is rare and restricted to outbreaks, exposure to the seasonal human coronaviruses is widespread. Population sero-surveys have shown that detectable baseline levels of IgG against at least one of the four known HCoV is near universal^12, 13, 14^, but there is evidence that re-infection with the same virus can occur^15, 16^. T cell immunity to other coronaviruses is less well studied prior to the 2020 pandemic, but a recent study from Singapore demonstrated the presence of reactive responses to SARS-CoV-2 in people who had recovered from the SARS-CoV epidemic 17 years earlier, which are likely to represent cross-reactive memory^9^. Such cross-reactive responses to HCoV may be protective against SARS-CoV-2, be irrelevant, or could in theory contribute to immunopathology. The role of pre-existing cross-reactive T cell responses in immunity has been studied for other viruses including flaviviruses. In one study where such responses were fine-mapped, we observed that pre-existing cross-reactive responses to dengue virus were linked to disease protection from Japanese Encephalitis, while symptomatic disease was linked to the emergence of strain-specific T cells^17^.

Divergent data regarding SARS-CoV-2 T cell cross-reactivity have emerged so far: recent studies of T cell immunity to SARS-CoV-2 have reported levels of cross-reactive immunity to HCoV in SARS-CoV-2 unexposed populations of between 0-50%^9, 10, 11, 18, 19, 20, 21^ using a variety of immune assays. One such study from our centre^20^ did not find significant *ex vivo* IFN-γ ELISpot responses to SARS-CoV-2 in uninfected, seronegative subjects. The differences between these results might reflect the use of different assays employing a range of antigenic targets, peptide concentrations and proliferation times.

Here we set out to address two questions using a panel of T cell assays. First, do COVID-19 patients and seronegative controls show different levels of responsiveness in distinct assays of T cell function? Second, can T-cell responses distinguish persons previously infected by SARS-CoV-2 from those previously infected by seasonal coronaviruses? We find - in a large cohort of subjects with a range of viral exposures – that cross-reactive memory responses to spike protein are almost universally detected using more sensitive assays, but that increasing viral exposure leads to an increase in magnitude and breadth of both effector and memory responses. These data have implications for our understanding of T cell cross-protection and for future studies of memory following the pandemic.

## Results

### High magnitude and broad IFN-γ T cell responses to SARS-CoV-2 structural proteins are present in convalescent subjects

We first examined the T cell response to SARS-CoV-2 in fresh PBMC using an *ex vivo* IFN-γ ELISpot assay from 168 subjects with PCR-confirmed SARS-CoV-2 infection, and 111 negative controls without evidence of SARS-CoV-2 infection (**Supplementary Table S1**). IgG antibody responses to spike measured by ELISA are shown in **Figure 1a** and neutralising antibodies measured by a pseudoparticle assay are shown in **Supplementary Figure 1a**. Firstly, we evaluated the magnitude of the T cell response to SARS-CoV-2 to assess the effector T cell response following stimulation of peripheral blood mononuclear cells (PBMCs) with pools of overlapping peptides spanning all SARS-CoV-2 proteins except the non-structural ORF1 (**Figure 1b and Supplementary Table S2**). We found responses to summed pools covering SARS-CoV-2 spike protein (12 mini-pools of 15-mers overlapping by 10 peptides referred to as P1 – P12) (**Figures 1b, 1c)**, and the structural and accessory proteins (7 pools of 18-mers overlapping by 11 peptides covering E, M, NP, ORF3, ORF6, ORF7 and ORF8) (**Figures 1b, 1d**). Median ELISpot responses to spike peptides were lower than those induced by immunisation with the candidate ChAdOx1^22^, although the vaccine trial ELISpot assay used a higher concentration of peptides. We also screened PBMCs with pools containing predicted optimal peptides targeting MHC Class II epitopes on the SARS-CoV-2 spike protein (CD4S), and other viral proteins (CD4All), and predicted Class I binding peptides split into CD8A and CD8B described in Grifoni et al, 2020^13^ (**Figure 1b, Supplementary Figure 1c**).

**Figure 1:**
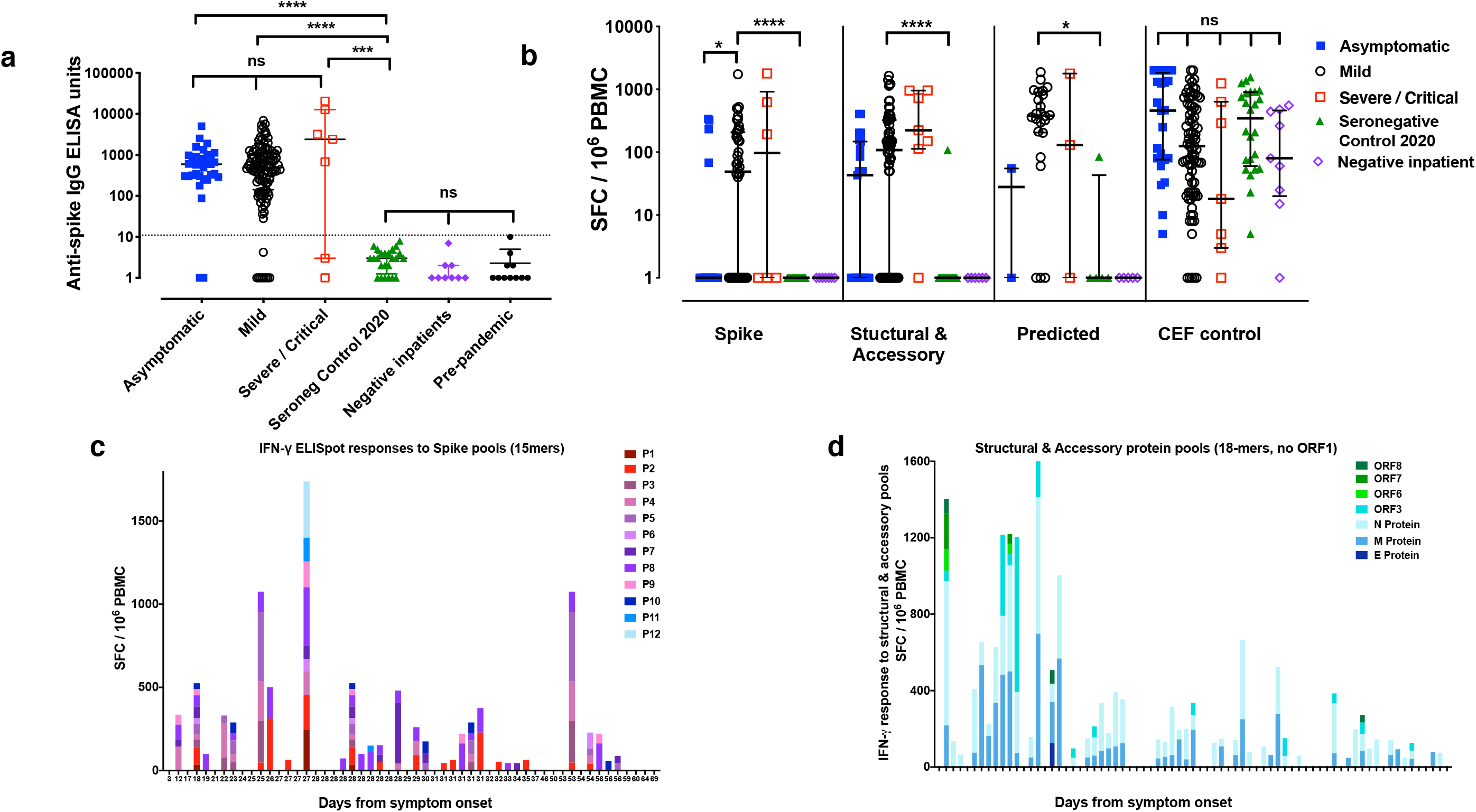
Magnitude and Breadth of SARS-CoV-2 specific Immune Response. **a)** Total anti-SARS CoV-2 spike IgG antibody titres by indirect ELISA^22^ in seronegative controls, asymptomatic and mildly symptomatic healthcare workers (HCWs) with PCR-confirmed SARS-CoV-2 infection, hospitalised patients with severe or critical PCR-confirmed SARS-CoV-2 infection, and PCR-negative inpatient controls, **b)** *Ex vivo* IFN-γ ELISpot showing the effector T cell responses to summed SARS-CoV-2 peptide pools spanning spike, accessory and structural proteins (E, M, N, ORF 3, ORF6, ORF7 and ORF8), *in silico* predicted pools^10^ and the CEF T cell control panel in subject groups as in a), **c)** *Ex vivo* IFN-γ ELISpot showing the magnitude and breadth of effector T cell responses to SARS-CoV-2 spike peptide pools and **d)** M, N, and accessory proteins ORF 3, ORF6, ORF7 and ORF8 in HCWs convalescent with mildly symptomatic SARS-CoV-2 infection. X axis shows number of days from onset of symptoms (not to scale), with blank columns representing zero response in the subject tested at that time-point. SFC/10^6^ PBMC = spot forming cells per million peripheral blood mononuclear cells, with background subtracted. ns = not significant, * = <0.05, ** = <0.01, *** = < 0.001 and **** = <0.0001, by Kruskal-Wallis one-way ANOVA, with Dunn’s multiple comparisons test for Figure 1b shown in Supplementary Table S5.

IFN-γ responses to spike (S) pools were seen in PBMC from 34/75 (45%) of convalescent subjects tested **(Figure 1c)** with notably high and frequent responses to the P2 (up to 313 SFC/10^6^ PBMC) and P8 minipools (up to 353 SFC/10^6^ PBMC). We identified IFN-γ responses to the structural and accessory proteins in 65/103 (63%) of convalescent subjects, with especially high-magnitude responses to the membrane (M) and nucleocapsid (NP) proteins **(Figure 1d)**. Combined, there was variation in the breadth and magnitude of SARS-CoV-2-specific responses **(Supplementary figure 1b)**, with an apparent peak of responses 28 – 32 days post onset of symptoms before declining, but longitudinal studies underway will better define the time course. IFN-γ responses were also seen in 24/29 (83%) of convalescent subjects following stimulation with the four pools of predicted epitopes. Interestingly, we found especially high-frequency responses to the CD8A pool which comprises predicted epitopes predominantly from the large ORF1^13,^ highlighting the need for further exploration of immune responses to this region **(Supplementary figure 1c)**.

### IFN-γ responses to either M or NP correlate with responses to different parts of the viral proteome

There was a correlation between summed responses to spike and non-spike structural proteins (Spearman R = 0.579, P < 0.0001, **Supplementary figure 1d)**, as well as the structural and accessory proteins and the predicted pools (data not shown), indicating that when an individual mounted a T cell response to one part of the proteome they were likely to respond to another part, and responses declined with time from symptoms **(Supplementary figure 1e and 1f)**. IFN-γ responses to either M or NP were correlates of the global response to spike, structural and accessory proteins **(Figure 2a and 2b)**, indicating that an assay to measure responses to M or NP could reflect the global effector T cell response.

**Figure 2:**
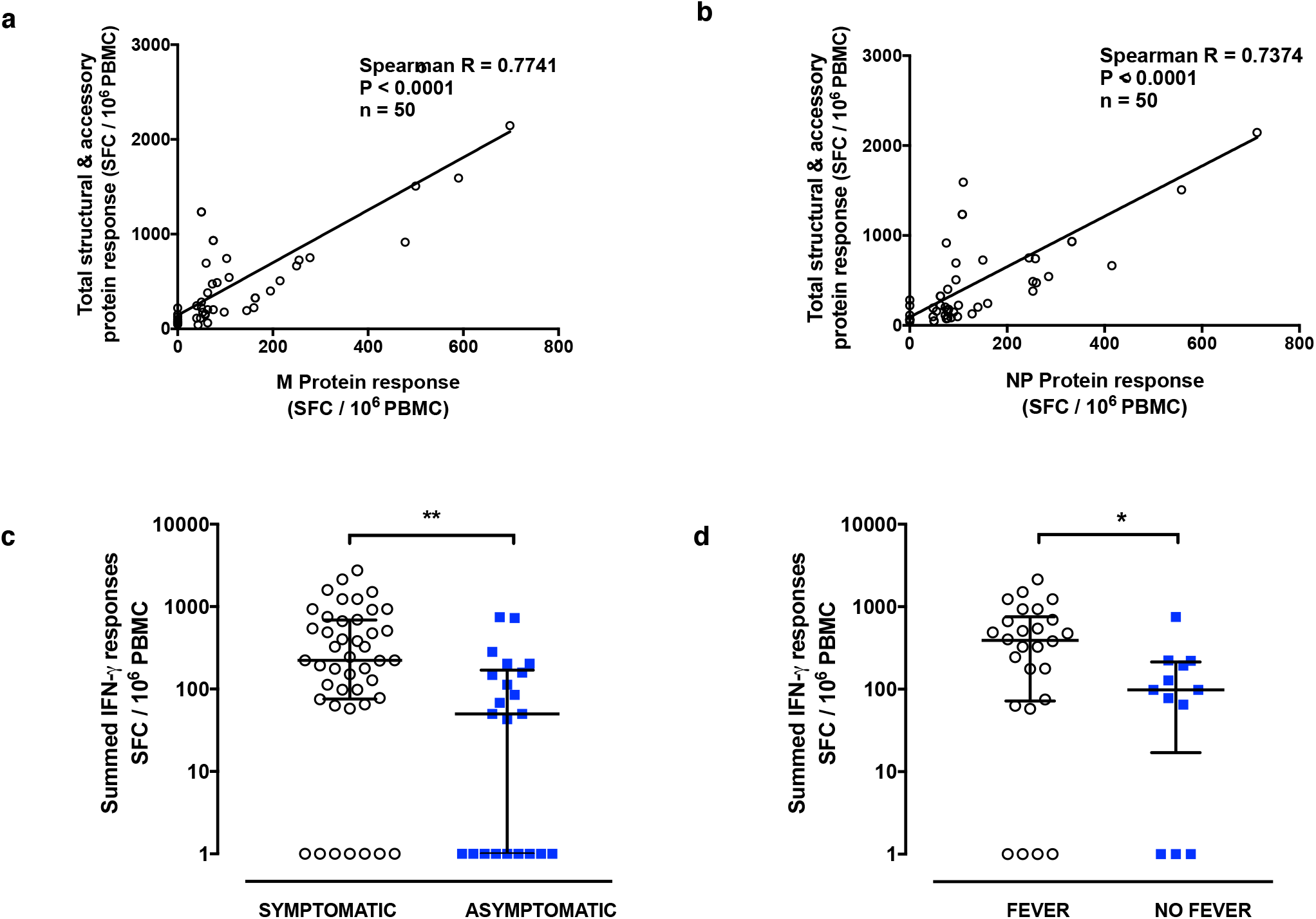
Total Summed *ex vivo* ELISpot Responses and Relationship with responses to individual protein responses and symptomology. Correlation between *ex vivo* IFN-γ ELISpot response to a) M protein and b) NP and total summed response to spike, E, M, N, ORF 3, ORF6, ORF7 and ORF8, c) Total summed response in HCWs with PCR-confirmed SARS-CoV-2 infection, with and without symptoms, d) Total summed response in the symptomatic HCWs who did and did not report fever. SFC/10^6^ PBMC = spot forming cells per million peripheral blood mononuclear cells, with background subtracted. Correlation was performed via Spearman’s rank correlation coefficient and comparison of two groups by Mann-Whitney U test. ns = not significant, * = <0.05, ** = <0.01, *** = < 0.001 and **** = <0.0001.

### Symptomatic SARS-CoV-2 infection and self-reported fever are associated with higher T cell responses

We found a significant difference (P=0.0013) in the magnitude of the IFN-γ response measured by ELISpot assay to spike and to the structural and accessory proteins depending on the presence or absence of symptoms (**Figure 2c and Supplementary Table S3**). It was not possible to explore the difference for the predicted optimal peptide pools due to insufficient numbers of asymptomatic subjects tested. Comparison between symptomatic and asymptomatic subjects may be confounded by differences in time from the start of the SARS-CoV-2 infection, which could be any time from 2 days to 6 weeks or more for the asymptomatic subjects^25^, and as demonstrated in **Supplementary Figure 1e and 1f**, time from infection impacts on the IFN-γ response. We therefore next examined whether a self-report of fever during COVID-19 in the symptomatic subjects was associated with a more vigorous T cell response by *ex vivo* IFN-γ ELISpot assay (P=0.0289, **Figure 2d and Supplementary Table S4**), and observed a greater magnitude of the IFN-γ response to spike in HCWs who reported fever compared to those who did not. There was no relationship for anti-spike IgG or neutralising antibody levels for the presence or absence of either symptoms or fever (data not shown), although another UK study has reported lower antibodies in asymptomatic and mild cases compared to more severe disease^8^. These results suggest that SARS-CoV-2 infections with a higher symptom burden such as fever induce a higher magnitude of T cell immunity than milder or asymptomatic infections. We did not find a significant difference between IFN-γ ELISpot response and either age or sex **(Supplementary Figures 1g and 1h)**, but larger studies including older adults are needed for further exploration.

### T cell proliferation assays are a sensitive method to demonstrate central memory responses to SARS-CoV-2

As our ELISpots assays were performed on total PBMCs, discrimination between distinct T cell lineages inducing the response was not possible. Moreover, the sensitivity of the ELISpot did not allow detection of responses in all COVID-19 recovered subjects. We therefore used a sensitive and functional flow cytometer-based assay capable of distinguishing the CD4+and CD8+ T cell responses. For this, we used a T cell proliferation assay to gain further insights into the contribution and relative proficiency of the CD4+ or CD8+ T cell compartments to drive a proliferative anti-SARS-CoV-2 immune response in our convalescent HCW cohort. We first validated our assays on a small cohort of healthy control subjects recruited for a hepatitis C virus (HCV) vaccine clinical trial pre-COVID19^23^. We showed that HCV seronegative control subjects made strong proliferative responses to pools of optimal peptides covering Influenza, EBV, CMV and Tetanus (FEC-T) but as expected, not to peptides covering HCV NS3 or core proteins **(Supplementary Figure S2a - S2e)**. We then evaluated the ability of CD4+ and CD8+ T cells from the COVID-19 convalescent HCW cohort to proliferate in response to peptide pools spanning key proteins from SARS-CoV-2. Live lymphocytes were separated into CD4+ or CD8+ T cells by gating strategy and the frequency of proliferating cells analysed following a 7-day stimulation **(Supplementary Figure S2a)**. We found a high frequency of proliferating cells and broad targeting of SARS-CoV-2-specific CD4+ and CD8+ T cells **(Figure 3a and 3b)** suggesting the establishment of a vigorous central memory population that may shape SARS-CoV-2 recall responses. The majority of subjects targeted T cell responses to M (69/107 for CD4+ and 50/107 for CD8+), NP (CD4+ 63/107; CD8+ 56/107) and ORF3 (CD4+ 26/107; CD8+ 24/107) and less frequently to ORF6 (CD4+ 4/107; CD8+ 2/107), ORF7 (CD4+ 11/107; CD8+ 6/107) and ORF8 (CD4+ 13/91; CD8+ 6/91) **(Figure 3c and 3d)**. This represents higher sensitivity to detect antigen-specific T cell responses than in the *ex vivo* ELISpot assay. Although we observed a trend for the overall magnitude of the proliferating CD4+ T cell response to SARS-CoV-2 peptide pools to be higher than that of the CD8+ T cell driven response, this did not reach significance for the peptide pools tested with the exception of M (P=0.0012) **(Supplementary Figure S3a**). Also of note, we did not find any difference in the magnitude of responding CD4+ or CD8+ T cells in individuals who had asymptomatic disease (detected on HCW screening) compared with those who presented with mild symptoms **(Supplementary Figure S3b and S3c)**. Finally, the findings from the proliferation assays were consistent with those generated by a second, shorter, assay measuring soluble lactate in supernatants obtained after only 4 days of stimulation, with SARS-CoV-2 convalescent subjects showing strong M, NP and ORF 3 directed responses **(Figure 3e)**. Taken together, in our cohort of convalescent HCWs we show wide breadth and magnitude of T cell responses to SARS-CoV-2 proteins including both subunits of spike (S1 and S2), and structural and accessory proteins.

**Figure 3:**
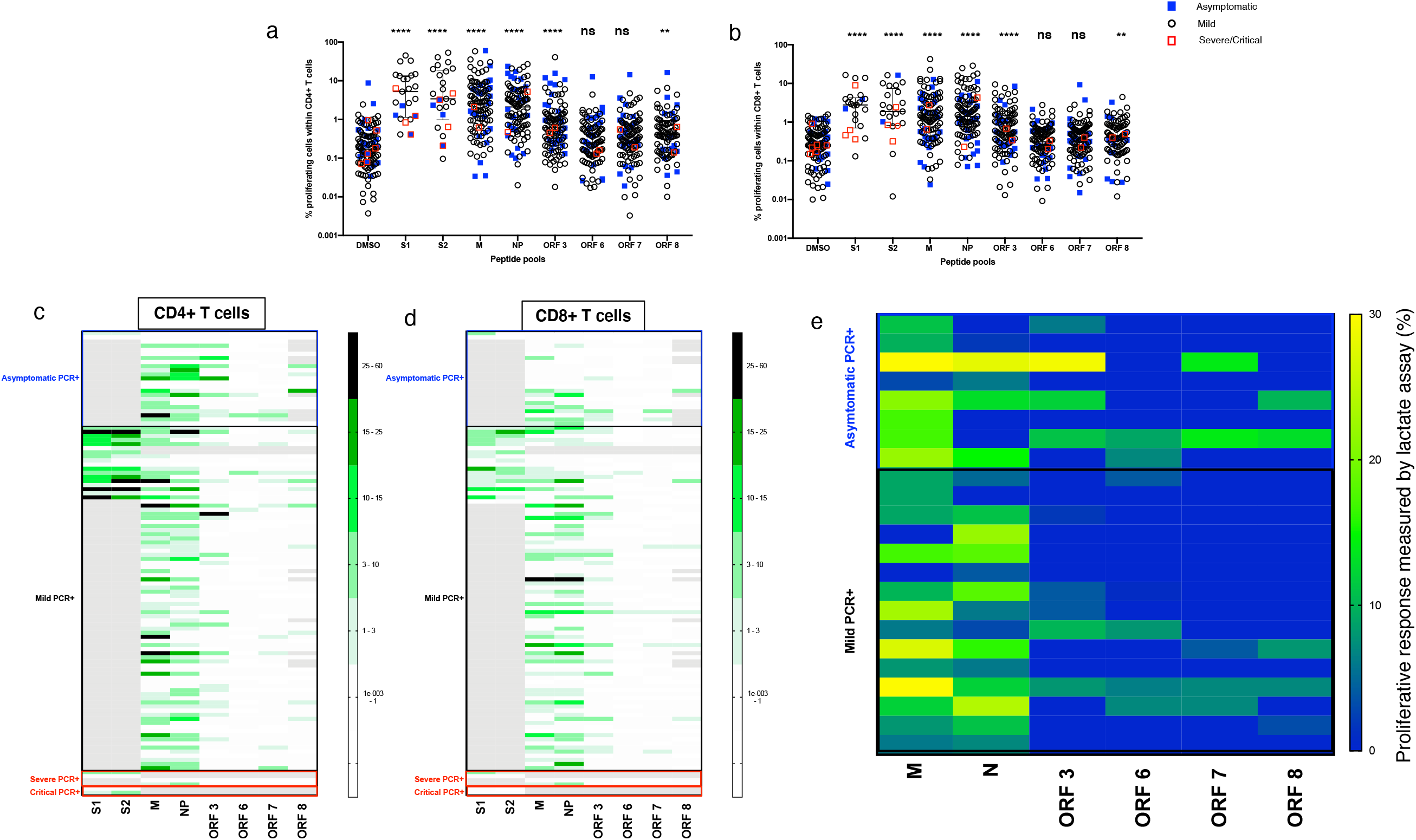
Proliferative responses in CD4+ and CD8+ T cells to key SARS-CoV-2 proteins. Plot showing raw frequency (without background subtraction) of proliferating cells in response to peptide pool stimulation in **a)** CD4+ and **b)** CD8+ T cells to DMSO (media), and overlapping peptide pools spanning S1, S2, M, NP, ORF 3, ORF6, ORF7, and ORF8. **c)** Heatmap showing magnitude of proliferative responses to overlapping peptide pools spanning SARS-CoV-2 proteome in CD4+ T cells and **d)** CD8+ T cells following background subtraction. Scales on heatmap represent magnitude of proliferating cells. Only datapoints >1% corresponding to mean + 2x SD in DMSO only well for both CD4+ and CD8+ T cells are shown. Grey box indicates absent data where tests were not run due to sample or peptide availability. **e)** Cellular lactate proliferative response in convalescent mild and asymptomatic HCWs at day 4 revealed a variable response to M, N, ORF 3, 6, 7, and 8. Heatmaps show background subtracted responses. Data shows media with interquartile range. ns = not significant, * = <0.05, ** = <0,01, *** = < 0.001 and **** = <0.0001 by Kruskal-Wallis one-way ANOVA, with Dunn’s multiple comparisons test for shown in Supplementary Tables S6a and 6b.

### T cell responses to SARS-CoV-2 in convalescent subjects are polyfunctional with both CD4+ and CD8+ responses

In order to determine the quality of the T cell response within our cohort we used an intracellular staining (ICS) panel comprised of the activation marker CD154, degranulation marker CD107a and effector cytokines IFN-γ, TNF-α and IL-2 on freshly isolated PBMC. This allows for the assessment of both the contribution of CD4+ and CD8+ T cell responses as well as the pattern of cytokine response to SARS-CoV-2. As both *ex vivo* ELISpot and ICS assays measure effector memory cells, we first focused our ICS analysis on subjects who were ELISpot responders (> mean + 2SD of the background) for M and NP pools, n=31 and 41 respectively. Representative plots are shown in **Supplementary Figure S4**. Levels of IFN-γ, IL-2 and TNF-α for these individuals are shown in **Figure 4a and 4d**. For M pools, there was a larger CD4+ T cell response compared to a CD8+ response in terms of both IL-2 (P<0.0001) and TNF-a (P=0.031) **(Figure 4a)**. Both cell types exhibited polyfunctionality with the majority of cells expressing one or two functional markers and up to four markers in CD4+T cells **(Figure 4b and 4c)**. For NP pools, there was no difference in the levels of IFN-γ, IL-2 or TNF-α expressed by CD4+ and CD8+ T cells (**Figure 4d**). This difference between M and NP responses was not due to differences in the magnitude of the ELISpot response as they were statistically similar (median 85 vs 95 SFC/10^6^ PBMC, *P*=0.37). Neither was this difference due to patients with asymptomatic disease as there were similar numbers who were ELISpot positive for M (n=8) and N (n=7). NP pools also stimulated a polyfunctional immune response with similar pattern as M pools **(Figure 4e, 4f)**

**Figure 4:**
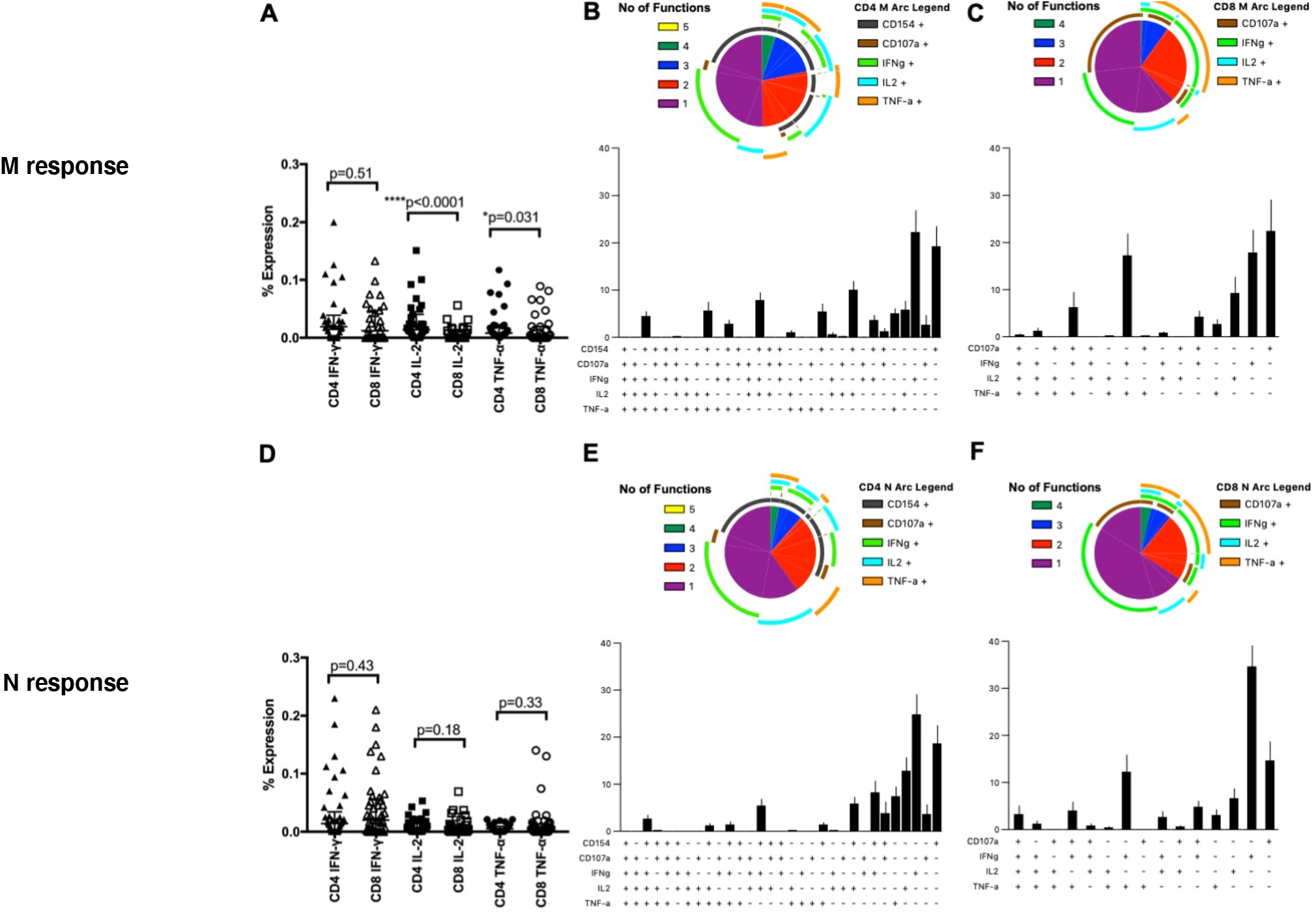
ICS responses in CD4+ and CD8+ T cells for M and NP pools in ELISpot positive individuals. ICS was performed on individuals with convalescent mild cases and a positive ELISpot for the indicated peptides. PBMC were stimulated with 2ug/mL peptide for 6 hours. Expression levels of IFN-γ, IL-2 and TNF-α in CD4+and CD8+T cells using M pools are shown in **a)**, n=31. Bars represent median +/- IQR. Statistics were performed using Wilcoxon matched-pairs signed rank test between each cytokine in CD4+vs CD8+T cells. Boolean gates were then set and polyfunctionality was examined in both CD4+(**b**) and CD8+T cells (**c**) using SPICE. For CD8+T cells, CD107a, IFN-γ, IL-2 and TNF-α were examined. CD154 was also analysed in CD4+T cells. Error bars represent SEM for polyfunctionality figures. Expression levels of cytokines using NP pools are shown in **d**) (n=41) with polyfunctionality analysis for CD4+T cells (**e**) and CD8+T cells (**f**) as above.

We then performed ICS experiments using M, NP, S1, and S2 pools on an additional 26 SARS-CoV-2 PCR positive individuals to compare the immune responses among these peptide pools (**Supplementary Figure S5**). M, S1, and S2 pools all trended towards higher levels of IL-2 expression by CD4+ T cells compared to CD8+ T cells (**Figure S5a, S5g, and S5j**, P=0.055, P=0.016, P=0.051, respectively). Stimulation with M pools also resulted in significantly higher expression of IFN-γ by CD4+ T cells (**Supplementary Figure S5a**, P=0.044) while NP pools trended towards higher IFN-γ expression in CD8+ T cells (**Figure S5d**, P=0.066). The vast majority of T cells (both CD4+ and CD8+) expressed 1-2 functional markers (with similar patterns), with small populations of CD4+ T cells expressing 3 (**Supplementary Figure S5**). The reduced polyfunctionality seen in this patient cohort is likely due to the lack of enrichment of ELISpot positive individuals, although potential loss in polyfunctionality with time may be a possible contribution as these donors were farther from symptom onset as a group.

### Seronegative control subjects show strong CD4+ and CD8+ T cell memory responses to the S1 and S2 subunits of the SARS-CoV-2 spike protein

We studied SARS-CoV-2 seronegative controls **(Figure 1a)** for whom we also evaluated T cell responses to SARS-CoV-2 peptides using IFN-γ ELISpot, ICS and proliferation assay. In contrast to convalescent HCWs, SARS-CoV-2-specific IFN-γ responses were scarcely seen in any of the SARS-CoV-2 peptide pools as measured by *ex vivo* ELISpot assays in 23 seronegative healthy control subjects **(Figure 1b)** and ICS (data not shown). Responsiveness to common antigens (CEF-T) in these control subjects indicated that there were no inherent defects in the ability of PBMCs from these donors to mount an antigen driven immune response. This finding of a lack of response to SARS-CoV-2 peptides in seronegative control subjects by an 18-hour *ex vivo* IFN-γ ELISpot assay was confirmed in 13 subjects by an independent laboratory in Sheffield, UK **(Figure 5a)**. We also evaluated cryopreserved PBMC from pre-pandemic healthy subject archives, and found minimal responses to spike, structural and accessory proteins in 12 subjects in Oxford (**Figure 5b**) and in the predicted epitope pools^10^ in 48 subjects in Liverpool, UK (**Figure 5c**).

**Figure 5:**
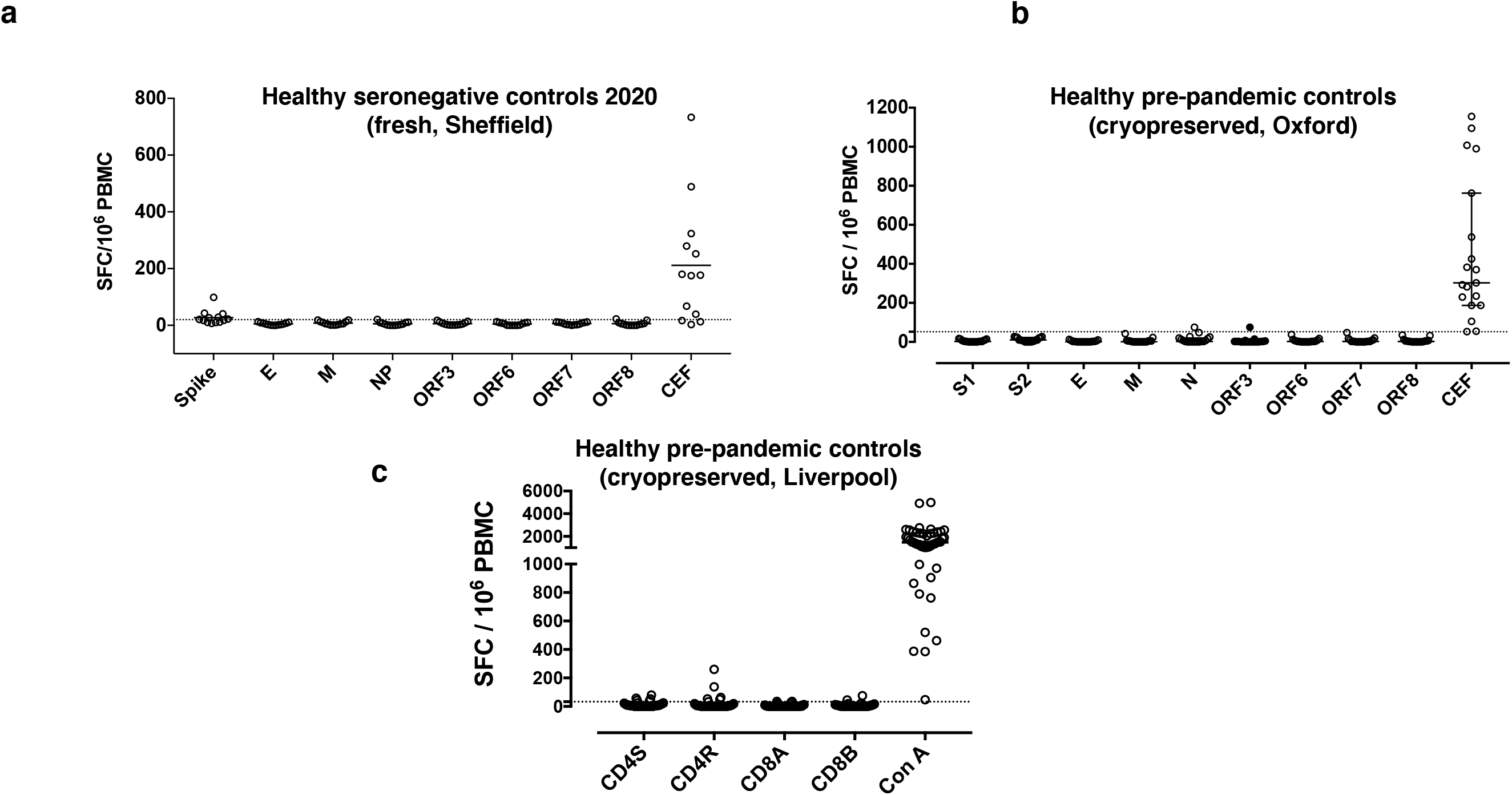
*ex vivo* ELISpot Responses in Seronegative Controls. *Ex vivo* IFN-γ ELISpot responses to summed SARS-CoV-2 peptide pools spanning spike, accessory and structural proteins (E, M, N, ORF 3, ORF6, ORF7 and ORF8) and CEF T cell control panel in a) freshly isolated peripheral blood mononuclear cells (PBMC) from seronegative controls in Sheffield, UK and b) cryopreserved PBMC from pre-pandemic healthy controls in Oxford. c) *Ex vivo* IFN-γ ELISpot responses to *in silico* predicted epitope pools^10^ cryopreserved PBMC from pre-pandemic healthy controls in Liverpool. Responses are shown with background subtracted, line represents mean + 2 stand deviations of responses to background.

However, using cellular proliferation assays on 20 seronegative subjects, we show high frequency of proliferating CD4+ and CD8+ T cells responding to the S1 and S2 subunit of the spike protein with a CD4+ T cell response detected in 17/20 (85%) and a CD8+ T cell response in 10/20 (50%) **(Figure 6a and 6b)**. In contrast, we observed weak or no CD4+ and CD8+ T cell proliferative responses to the structural and accessory proteins studied (M, NP, ORF3, ORF6, ORF7 and ORF8 **(Figure 6a and 6b)**. As the 20 seronegative participants were sampled in early 2020, we also analysed cryopreserved samples from 2008-2019 (pre-UK COVID19 pandemic) to exclude the possibility of asymptomatic and undetected prior infection. Similar to the pandemic seronegative controls, we found no or low effector T cell responses by ELISpot assay to any of the spike, structural or accessory proteins (**Figure 5b**), but as for the pandemic seronegative controls we detected robust T cell responses by proliferation assay to spike proteins S1 and S2 which was of greater breadth in the CD4+ T cells compared to their CD8+ T cell counterparts **(Figure 6c and 6d)**. The responses show a CD4+ skew with 15/15 showing a CD4+ T cell response and only 8/15 showing a CD8+ T cell response above background level. Most importantly, there was very limited cross-reactivity to the structural and accessory proteins as measured by the proliferation assay. As with the convalescent HCW cohort, we also performed a cellular lactate assay using supernatants obtained after 4 days of stimulation on 8 of these subjects. We confirm cross-reactive responses to spike S1 and S2 subunits, and non-existent or minimal responses in supernatants obtained from M, NP and accessory protein-stimulated PBMCs **(Figure 6e)**. We compared the magnitude of the proliferative responses to the different SARS-COV-2 peptide pools in seronegative controls from 2020, symptomatic and asymptomatic SARS-COV-2 PCR+ subjects **(Figure 6f and g)**. We found no difference in the spike – S1 and S2 - responses but higher magnitude of proliferative responses to M and NP in both CD4+ and CD8+ T cells and ORF3 and ORF8 in CD8+ T cells alone in subjects who had tested positive to SARS-COV-2 **(Figure 6f and g)**. For confirmation, we also compared the magnitude of proliferative T cell responses in SARS-COV-2 seronegative controls from 2020 with the cryopreserved pre-pandemic seronegative controls and found the magnitude of proliferative cells in these two seronegative groups to be similar **(Supplementary Figure S6a and S6b)**. These results, in addition to our earlier results from subjects who did not generate effector T cell responses to spike peptides in the IFN-γ ELISpot assay **(Figure 1b and Figures 5a-5c)**, demonstrate the existence of central memory T cell immunity to spike protein in the pre-existing T cell repertoire of subjects naïve to SARS-CoV-2.

**Figure 6:**
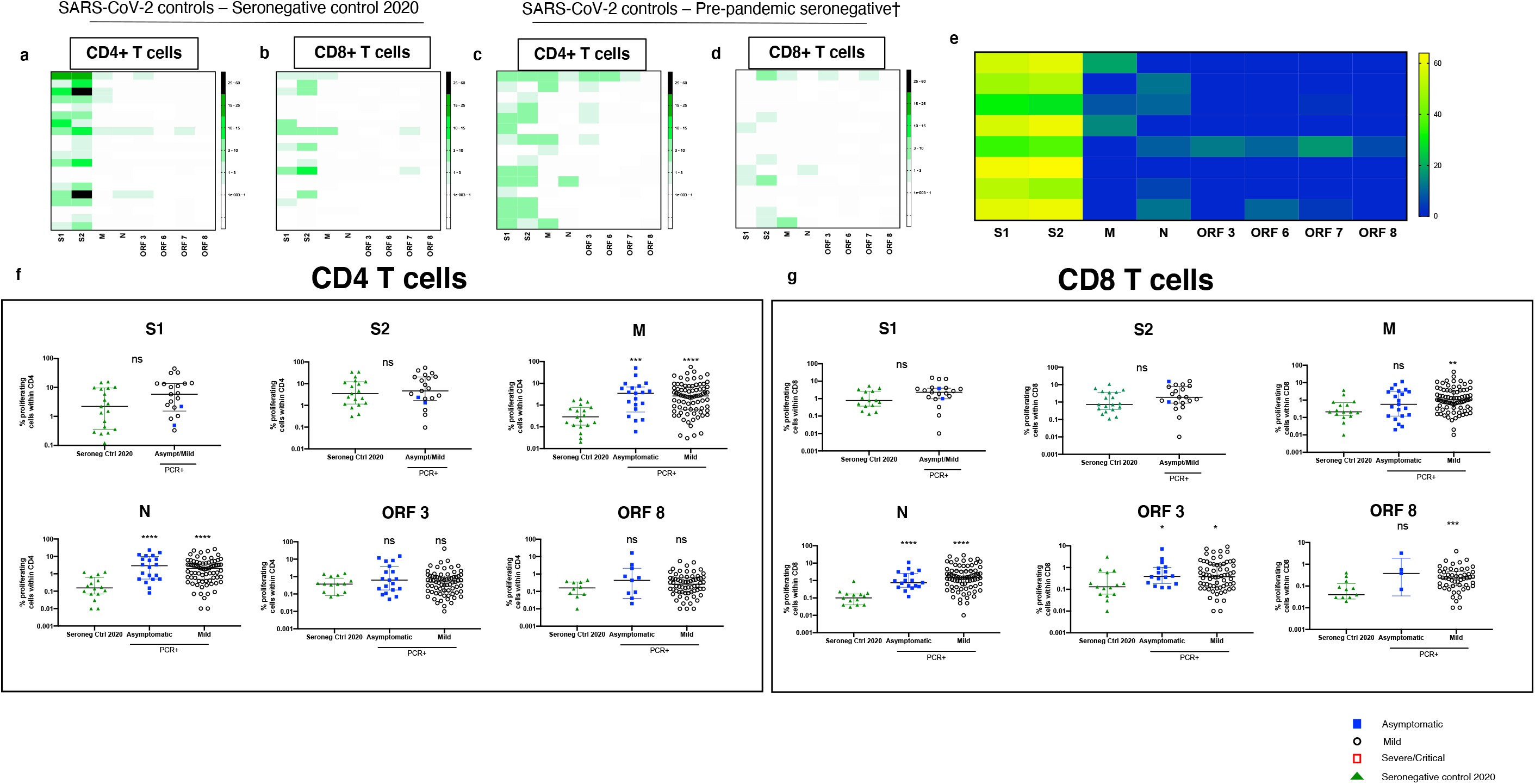
Cross-reactive T cell response in Seronegative Controls. **a)** Heatmaps showing CD4+ and **b)** CD8+ T cell proliferative responses in fresh PBMCs from healthy seronegative controls. **c)** heatmaps showing the magnitude of cross-reactive responses in CD4+ and **d)** CD8+ T cell response in cryopreserved samples obtained pre-COVID19 pandemic. Only datapoints >1% corresponding to mean + 2x SD in DMSO only well for both CD4+ and CD8+ T cells are shown. Grey box indicates absent data where tests were not run due to sample or peptide availability. **e)** Heatmap measuring the lactate proliferative response in both healthy seronegative controls at day 4 revealed a strong response to spike (all S1 and S2 values divided by 2 for ease of viewing) as well a small, variable, response to SARS-CoV-2 peptide pools. **f)** comparative analysis of peptide pool specific proliferative response to SARS-CoV-2 proteins in CD4+ and **g)** CD8+ T cells in SARS-CoV-2 seronegative controls during COVID pandemic and PCR+ volunteers. All data plotted are background subtracted. For statistical comparison, all datapoints have been included for all groups. Data shows media with interquartile range ns = not significant, * = <0.05, ** = <0,01, *** = < 0.001 and **** = <0.0001. Dotted lines on plot indicate 2.5x DMSO background analysed on a cross-sectional level

### Application of T-cell assays reveal responses in seronegative highly exposed healthcare workers

Finally, to explore the use of these T cells assays to identify people potentially exposed to SARS CoV-2, we recruited a group of 10 highly exposed doctors working in acute medicine who had experienced symptoms compatible with COVID-19 but had not received PCR testing at the time of symptoms, or tested negative, and were subsequently seronegative. 3/10 of these subjects showed effector T cell responses by ELISpot assay to S1, S2, M or NP **(Figure 7a)** whilst 8/8 of those tested showed M and / or NP-specific T cell responses in the proliferation assay compatible with prior infection **(Figure 7b and 7c)**. For one donor where cells were available, a fresh ICS assay confirmed this ELISpot response (data not shown). Analysis of the breadth of SARS-CoV-2 antigen targeted by the responding CD4+ and CD8+ T cells shows that in the highly exposed doctors the CD4+ and CD8+ T cell response is directed to a broader number of structural (M and NP) and accessory (ORFs 3, 6, 7 and 8) SARS-CoV-2 peptide pools **(Figure 7d and e)**. This reached statistical significance for both CD4+ and CD8+ T cells compared to seronegative control groups **(Figure 7d and e)**. Lastly, we compared the magnitude of the T cell response to SARS-CoV-2 structural and accessory proteins in the three groups – the highly exposed doctors, seronegative controls from 2020 and pre-pandemic seronegative controls (combined into one group). We found significantly higher magnitude of CD4+ but not CD8+ T cells proliferating in response to the M, N, ORF3, 6, 7 and 8 in the highly exposed doctors **(Figure 7f and g)**.

**Figure 7:**
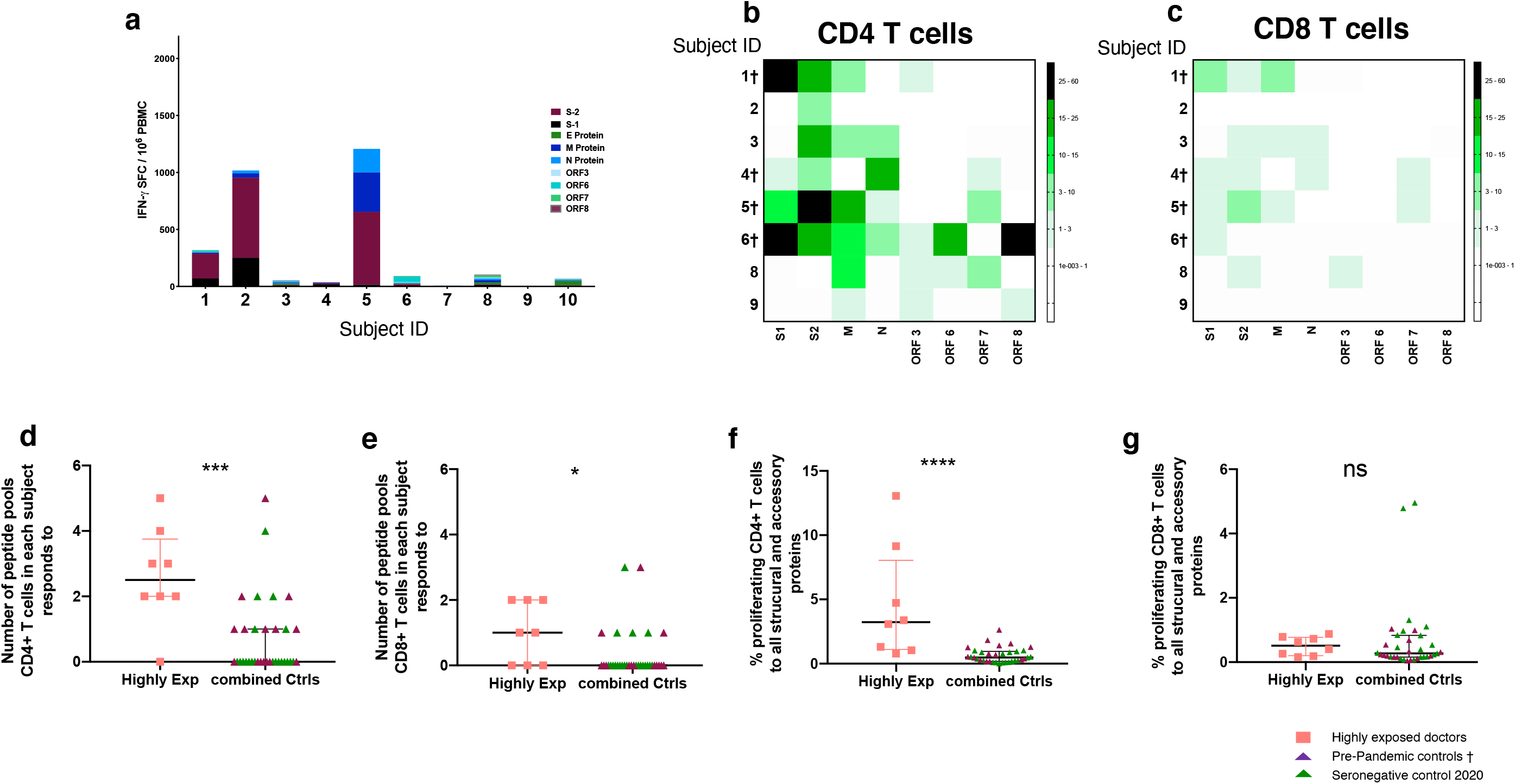
T cell Response in Highly Exposed Seronegative Controls. *Ex vivo* IFN-γ ELISpot responses to summed SARS-CoV-2 peptide pools spanning spike, accessory and structural proteins (E, M, N, ORF 3, ORF6, ORF7 and ORF8) and CEF T cell control panel in highly exposed doctors working in acute medical care who experienced a COVID-19 compatible illness without PCR testing and were subsequently seronegative. Responses are shown with background subtracted. **b)** Heatmaps showing CD4+ and **c)** CD8+ T cell proliferative responses in the same population of highly exposed doctors. All data plotted are background subtracted. **d)** Breadth of responses to structural and accessory proteins from SARS-COV-2 in CD4+ and **e)** CD8+ proliferating T cells. **f)** Magnitude of responding CD4+ and **g)** CD8+ T cells to structural and accessory proteins from SARS-CoV-2 (M, N, ORF3, 6, 7, 8). Subject ID with † were assessed from cryopreserved samples. Proliferation assay for subjects 7 and 10 was not performed.

## Discussion

As the global COVID-19 pandemic continues, it is important to define which immune responses are important for protection. In this study we have used distinct T cell assay platforms across the same individuals to identify the differences between T cell responses associated with recent SARS-CoV-2 infection and long-term cross-reactive memory T cell responses in unexposed populations. The effector T cell response as measured by our 18-hour *ex vivo* IFN-γ ELISpot assay showed a remarkable absence of SARS-CoV-2 specific responses in most of the healthy seronegative subjects. The ELISpot assay therefore represents potential as a specific assay for identification of recent infection with SARS-CoV-2, although the longevity of such responses requires further analysis in longitudinal studies. We already noted a significant inverse correlation with magnitude by ELISpot assay over time in the short follow up performed here, and ongoing work with the current convalescent HCW cohort will define the durability of these T cell responses induced by SARS-CoV-2 infection.

In contrast, the same healthy subjects showed responses to the S1 and S2 subunits of spike protein in a 7-day CTV proliferation assay, confirmed by analysis of lactate production. The most likely explanation for this is that people retained cross-reactive central memory responses to the spike protein of seasonal coronaviruses that circulate in the UK, although cross-reactivity from other human micro-organisms is also possible, as has been described for HIV, influenza and Ebola epitopes in naïve subjects^24, 25^. These cross-reactive responses may have been underestimated in previous reports using more *ex vivo* type assays with limited sensitivity.

Individuals in convalescence from SARS-CoV-2 infection showed strong and broad effector CD4+ and CD8+ polyfunctional T cell responses to peptides spanning the SARS-CoV-2 genome as previously reported^20^. ELISpot responses to the M and NP proteins were especially frequent and high, and each correlated with the summed response to spike, structural and accessory peptides, indicating their suitability as antigens for screening individuals and populations for evidence of T cell immunity following exposure to SARS-CoV-2. Additionally, central memory responses to M and NP were frequent and strong in the proliferation assay for subjects in convalescence from SARS-CoV-2 but significantly less so in the seronegative control subjects, further supporting the use of these antigens as markers of T cell responsiveness more closely linked with SARS-CoV-2 exposure. Further mapping studies could identify peptides with the highest sensitivity and specificity for SARS-CoV-2 infection, with potential for use in defining T cell immunity at an individual and population level.

The existence of substantial T cell cross-reactivity to SARS-CoV-2 from prior HCoV exposure has been demonstrated in non-SARS-CoV-2 infected populations from a range of geographical locations ^9, 10, 11, 18, 19, 21^. Here, we demonstrate use of the ELISpot assay to identify SARS-CoV-2 specific responses, and our finding of absent T cell responses in unexposed subjects was confirmed by similar results in our three independent laboratories (Universities of Oxford, Liverpool and Sheffield). T cell assays vary in their sensitivity, influenced by cell number, incubation time, antigen choice and concentration and markers of T cell activity measured. Our ELISpot assay does not detect the T cell responses in unexposed populations to spike and other SARS-CoV-2 proteins reported elsewhere. This may be due to the relatively low cell number used in our assay (200,000 per well) but most likely the focus on IFN-γ release rather than detection of cell activation markers. In this data we see a greater magnitude of ELISpot responses in convalescent symptomatic subjects who reported fever during their illness, compared with symptomatic subjects who did not report fever. We also saw higher responses in symptomatic people compared with asymptomatic people. Febrile symptomatic disease represents a greater systemic response and such individuals appear to mount a more vigorous T cell response (akin to a higher vaccine dose). This could represent failure of early/innate immune control necessitating a larger adaptive response and that hypothesis is consistent with the correlation we saw between T cell ELISpot and ELISA antibody levels. This is seen in other settings – for example, higher antigen-specific CD4+ T cell responses in more severe cases of H1N1/09 influenza A^26^.

Most convalescent subjects in the study made antibodies, as detected by IgG ELISA and pseudoparticle neutralisation assay. Emerging literature suggests that SARS-CoV-2 IgG titres meeting the threshold for positivity may be relatively short-lived^8, 27^. The current study represents a cross-sectional “snapshot” in time of human T cell responses to SARS-CoV-2 after infection, and the data suggests a peak in magnitude of the effector T cell response around 4 weeks after the onset of symptoms. Ongoing follow-up studies of this cohort and surveillance^28^ for re-infection aligned to the UK SIREN study^29^ will allow further delineation of the time course of T cell responses in parallel with humoral responses, and the timing of any assay must be taken into account in defining its utility. While an association is seen between antibody and ELISpot in the PCR-positive cohort, a disjunct exists between the antibodies and memory responses, since strong spike responses can be seen in the PCR-negative / unexposed and pre-pandemic groups. We need to assess in future whether any relationship exists between the levels of these responses and levels of seroreactivity to HCoVs.

Our study of the large ORF1 was restricted to use of the *in silico* predicted pool CD8A^10^, where high magnitude of responses were seen. Further work will characterise the time course of T cell responses observed in this cohort, evaluate the ability of our assays to correctly distinguish individuals with confirmed SARS-CoV-2 infection from unexposed controls, and prospectively seek to identify the relationship between measurable T cell immunity to the SARS-CoV-2 proteome and subsequent primary or secondary infection with SARS-CoV-2.

Overall, we have shown that assessments of T cell immunity using different assays but with the same antigens give very different results. Our ELISpot measure of *ex vivo* IFN-γ release is valuable in defining the potential role of T cell immunity in recently infected donors without cross-reactivity in unexposed subjects. In contrast, our proliferation assay allows dissecting out pre-existing vs SARS-CoV-2 induced immune responses by examining responses to different antigens. Our proliferation assay demonstrates widespread T cell memory responses to spike in both SARS-CoV-2 infected and unexposed subjects, whilst T cell memory responses to M and NP are characteristic of previous SARS-CoV-2 infection. These two assays, in combination with the panel of antigens can now allow us to address critical questions about the role of T cells – induced by SARS-CoV-2, by HCoVs or by vaccines - in immune protection in the future.

## MATERIALS AND METHODS

### Ethics Statement

Human study protocols were approved by the research ethics committee (REC) at Yorkshire & The Humber - Sheffield (GI Biobank Study 16/YH/0247). The study was conducted according to the principles of the Declaration of Helsinki (2008) and the International Conference on Harmonization (ICH) Good Clinical Practice (GCP) guidelines. Written informed consent was obtained for all patients enrolled in the study.

### Study subjects

#### (i) SARS-CoV-2 positive individuals

Healthcare workers at Oxford University Hospitals NHS Foundation Trust who tested positive for SARS-CoV-2 following either presentation to the hospital’s Occupational Health Department with symptoms or had a positive PCR test on the staff screening programme^28^ were asked to indicate whether they were willing to be contacted by researchers. Individuals who agreed to be contacted received an email invitation to participate in the study. Subjects recruited from the staff screening programme were classified as asymptomatic if they did not report any symptoms of COVID-19 (including fever, shortness of breath, cough, loss of taste or smell, sore throat, coryza or diarrhoea), either prior to staff screening or in the seven days following testing positive. In total 126 symptomatic and 33 asymptomatic subjects were recruited for this study. In addition, 9 hospitalised patients with WHO severe or critical COVID-19 were studied.

#### (ii) SARS-CoV-2 negative individuals (healthy controls)

30 healthy control subjects in Oxford and 13 in Sheffield with no history of COVID-19 symptoms and no antibodies to SARS-CoV-2 spike protein detected by IgG ELISA were recruited. In addition, archived samples from 12 healthy control subjects in Oxford who donated blood in the pre-pandemic period (2008-2019) were studied, alongside 48 healthy control subjects from the pre-pandemic period in Liverpool. Hospital in-patients with PCR-confirmed SARS-CoV-2 infection or PCR-negative inpatient control subjects were recruited by the study team.

#### (iii) Highly exposed seronegative individuals (highly exposed)

10 acute medicine doctors, who worked in patient facing services during the pandemic and experienced symptoms compatible with COVID-19, but did not receive PCR testing at the time of symptoms or tested negative, and were anti-spike IgG negative two months after the pandemic peak, were recruited as highly exposed seronegative participants.

### Peripheral blood mononuclear cells

Peripheral blood mononuclear cells were isolated by density gradient centrifugation using Lymphoprep™ (p=1.077 g/ml, Stem Cell Technologies) as previously described^30^. Plasma was collected and spun at 2000g for 10 minutes to remove platelets before freezing at −80°C for later use. PBMC were collected and washed twice with pre-warmed R10 media: RPMI 1640 (Sigma, St. Louis, MO, USA) supplemented with 10% heat-inactivated FCS (Sigma), 1mM Pen/Strep and 2mM L-Glutamine (both from Sigma). After the second centrifugation, cells were resuspended in R10 and counted using the Guava® ViaCount™ assay on the Muse Cell Analyzer (Luminex Cooperation). The majority of assays were performed on freshly isolated PBMC during the first peak of the pandemic using available resources, and it was not possible to test all samples with all antigens. Assays performed on frozen samples are indicated in the manuscript.

### Antigens

For functional assays, PBMC were stimulated with three groups of peptide pool for SARS-CoV-2: (1) Spike: 15-mers overlapping by 10 amino acid residues for spike (S), divided into 12 “minipools” P1-P12 (Proimmune)^22^, and grouped into pools S1 (P1-6) and S2 (P7-12) for some assays (2) Structural and accessory proteins: 12-20-mer peptides overlapping by 10 amino acid residues for membrane protein (M), nucleoprotein (NP), envelope (E) protein, open reading frame (ORF) 3, 6, 7 and 8 (Proimmune)^20^ and (3) Predicted epitope pools: predicted CD4+ and CD8+ pools^10^ from the Sette laboratory, La Jolla Institute, CA, all used at a final concentration of 1-2ug/ml per peptide depending on the assay. Lyophilised peptides were reconstituted in DMSO (Sigma) A list of peptide sequences and composition of peptide pools is provided in **Supplementary Table S5**.

### IFN-γ enzyme-linked immunospot (ELISpot) assay

The kinetics and magnitude of the cellular responses to SARS-CoV-2 were assessed by *ex-vivo* IFN-γ ELISpot as previously described^30^. Fresh PBMC were used in all ELISpot assays unless otherwise indicated in figure legends. Briefly, 96-well Multiscreen-I plates (Millipore, UK) were coated for 3 hours with 10 μg/ml GZ-4 anti-human IFN-γ (Mabtech, AB, Sweden) at room temperature. Fresh PBMC were added in duplicate wells at 2×10^5^ cells in 50 μl per well and stimulated with 50 μl of SARS-CoV-2 peptide pools (2ug/ml per peptide) as indicated in the figure legends and controls. R10 with DMSO (final concentration 0.4%, Sigma) was used as negative control and the following reagents were used as positive controls: CEFT peptide pool (2µg/ml, Proimmune) and Concanavalin A (5µg/ml final concentration, Sigma). After 16-18 hours at 37°C, 5% CO2, 95% humidity, cells were removed and secreted IFN-γ was detected by adding 1 μg/ml anti-IFN-γ biotinylated mAb (7-B6-1-biotin, Mabtech) for 2-3 hours, followed by 1 μg/ml streptavidin alkaline phosphatase for 1-2 hours (SP-3020, Vector Labs). The plates were developed using BCIP/NBT substrate (Pierce) according to the manufacturer’s instructions. ELISpot plates were scanned on an AID ELISpot Reader (v.4.0) using the following settings: Intensity min 12, Size min 22, Gradient min 4. Results were reported as spot-forming units (SFU) per million PBMC. The unspecific background (mean SFU from negative control wells) was always less than xx SFU/10^6^ PBMC and subtracted from experimental readings.

### Intracellular cytokine stimulation (ICS) assay

PBMC resuspended in R10 were plated at 1×10^6^ live cells/well into 96 well round bottom plates and stimulated with SARS-CoV-2 peptide pools (2ug/ml per peptide) as indicated in the figure legends. Media containing DMSO (0.1%, Sigma) was used as negative control and PMA (0.05ug/mL) with ionomycin (0.5ug/mL, Sigma) as a positive control. CD107a BV421 (BD Biosciences) and Brefeldin A (MP Biomedicals) were added to cultures at a final concentration of 0.04ug/mL and 10ug/mL respectively and cells were incubated for 6 hours at 37°C, 5% CO2, 95% humidity. Plates were placed at 4°C overnight and subjected to flow cytometry staining as described below. In addition to the three cytokines, CD107a was examined as was CD154 in CD4+ T cells.

### Proliferation assay

PBMCs from freshly isolated blood samples or cryopreserved samples (denoted with †) were twice washed with 1x PBS and stained using CellTrace® Violet (CTV, Life Technologies) at a final concentration of 2.5uM for 10 minutes at room temperature. The reaction was quenched by adding cold FBS. CTV-labelled PBMC in RPMI containing 10% human AB serum (Sigma), 1mM Pen/Strep and 2mM L-Glut were plated in a 48 or 96 well round bottom plates at 500,000 and 250,000 cells respectively and stimulated with peptide pools from SARS-CoV-2, FEC-T, HCV NS3 or HCV core protein (1μg/ml per peptide). Media containing 0.1% DMSO (Sigma) representing DMSO content in peptide pools was used as negative control and 2 μg/ml phytohemagglutinin L (PHA-L, Sigma) as used as positive control. Cells were subsequently incubated at 37°C, 5% CO2, 95% humidity for 5 days without media change or 7 days with media change on day 4 if cultures were kept beyond 5 days. At the end of incubation, cells were subjected to flow cytometry staining as described below. Responses above 1% were considered true positive. To determine the breadth of antigenic response targeted by T cells, the number of peptide pools that each subject responded to was counted. To determine the magnitude of the total response to structural and accessory proteins, the average number of cells proliferating in response to any of the peptides M, N, ORF3, 6, 7, 8 was obtained as a function of their respective CD4+ or CD8+ T cell population and then expressed as a percentage. Background was then subtracted from the total response for each subject.

### Flow cytometry staining

A MIFlowCyt file (minimum information about a flow cytometry experiment) was created as per Section 4 of “Guidelines for the use of flow cytometry and cell sorting in immunological studies”^31^ and recommended by the International Society for Advancement of Cytometry^32^. The file contains details of antibodies, reagents, instrument settings, gating strategies and controls used for flow cytometry experiments and is provided in the supplementary information of this manuscript. PBMC were resuspended in cell staining buffer (Biolegend) in case of proliferation assays or 1xPBS in case of ICS assays and incubated for 20min with near-infrared live/dead or aqua fixable stain, respectively (Invitrogen, Carlsbad, CA, USA). Cells from proliferation assays were incubated with fluorochrome-conjugated primary human-specific antibodies for CD3, CD4 and CD8 in cell staining buffer (Biolegend) containing serum for 30min at 4°C, washed with cell staining buffer, fixed with 4% paraformaldehyde (PFA, Sigma) and stored at 4°C in the dark until data acquisition. Cells from ICS assays were fixed with fixation/permeabilization solution (BD Biosciences) for 20 min at 4°C, washed with permeabilization buffer (BD Biosciences) followed by incubation with fluorochrome-conjugated human-specific antibodies. After washing with permeabilization buffer, the samples were resuspended in 1xPBS and stored at 4°C in the dark until data acquisition. Data was acquired on an LSRII flow cytometer (BD Biosciences) and analysis was performed with FlowJo Version 10 (BD Biosciences). Specific gating strategies can be found in the Supporting information (MIFlowCyt File).

### Lactate measurements

Supernatants from the proliferation assay were analysed using a previously published assay^33^. Briefly, colorimetric L-lactate assay kits (Abcam, Cambridge, UK) were used as per manufacturer’s instructions. A standard concentration curve was defined, and the lactate concentration in each day 4 supernatant from the proliferation assay was calculated using a 96-well plate reader.

The lactate proliferation index was calculated on a per-well basis using the following equation 1:

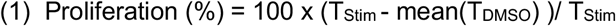

Where T_Stim_ is the concentration of lactate for a given well with either PHA or SARS-CoV-2 peptides, and mean(T_DMSO_) is the average background lactate production from negative control wells.

A significant proliferative response to a given peptide was greater than 0, as determined by equation 2:

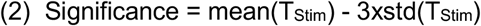

Where mean(T_Stim_) is the mean % proliferative response of a specific participant to a stimulus, and std(T_Stim_) is the standard deviation of the participant to a given stimulus.

### Standardised ELISA for detection of SARS-CoV-2 spike antigen-specific total IgG in plasma

Total anti-SARS CoV-2 spike antibodies were determined using an indirect ELISA as described previously^22^, which is based on the Krammer assay^34^ using a standard curve derived from a pool of SARS-COV-2 convalescent plasma samples on every plate. Standardised EUs were determined from a single dilution of each sample against the standard curve which was plotted using the 4-Parameter logistic model (Gen5 v3.09, BioTek). Each assay plate consisted of samples and controls plated in triplicate, with ten standard points in duplicate and four blank wells.

### SARS-CoV-2 pseudotype micro-neutralisation assay

Frozen plasma samples were thawed, heat-inactivated at 56C for 30 minutes, and assayed for neutralisation of a lentivirus-based viral particle carrying a luciferase reporter and pseudotyped with full-length SARS-CoV-2 spike (Accession No: YP_009724390.1), as detailed in Thompson et al^35^. Briefly, neutralising antibody titres were determined by incubating serial two-fold plasma dilutions with ∼10^5 RLU pseudotyped virus for 2hrs before addition of 10^4 HEK293T cells transfected with full-length human ACE2 24hrs prior. After 72hrs incubation at 37C, luciferase expression was quantified using BrightGlow (Promega Corp.), readouts were normalised, and -Log(IC50) determined via non-linear regression using GraphPad Prism8 (GraphPad Software).

### Statistical analyses

Statistical analysis was performed with IBM SPSS Statistics 25 and figures were made with GraphPad Prism 8. Chi-square was used to compare ratio difference between two groups. After testing for normality using Kolmogorov-Smirnov test, Independent-samples *t* test or Mann-Whitney U test was employed to compare variables between two groups, and Kruskal-Wallis-ANOVA with Dunn’s multiple comparisons test was performed to compare variables between three or more groups with a non-parametric distribution. Correlation was performed via Spearman’s rank correlation coefficient. For polyfunctionality analyses, data was prepared using PESTEL v2.0 for formatting and baseline subtraction, followed by export of data to SPICE v6.0 for analysis. Statistical significance was set at P<0.05 and all tests were 2-tailed.

## Data Availability

All data will be made public upon publication of the results in a peer-reviewed journal

## Acknowledgements

The authors wish to thank Kathryn Southey and Suki Kenth for administrative support, and our medical student volunteers Julie Dequaire, Rory Fairhead, Shayan Fassih, Thomas Foord, David Kim, Thomas Ritter, Adan Taylor, Rebecca Young for logistical assistance.

---

**Supplementary Table S1:** Clinical Information for Patients used in the Study.

| **Group** | **N** | **Sex**<br>**F/M (%)** | **Age**<br>**- median (IQR)** | **Time from PCR**<br>**- median (IQR)** | **Time from**<br>**symptoms -**<br>**median, (IQR)** |
| --- | --- | --- | --- | --- | --- |
| **Asymptomatic** | 33 | 26/7<br>(79/21%) | 36 (27-47) | 6 (5-28) | n/a |
| **Mild** | 126 | 92/34<br>(73/27%) | 34.5 (27-48) | 29 (24-54) | 34.5 (27-57) |
| **Severe & Critical** | 9 | 3/6<br>(33/67%) | 52 (41-69) | 18 (9-24) | 23 (13-29) |
| **TOTAL PCR-confirmed**<br>**SARS-CoV-2 infection** | 168 |  |  |  |  |
| **Healthy controls:**<br>**Contemporaneous**<br>**(Oxford)** | 30 | 13/17<br>(43/57%) | 37 (33-47) | n/a | n/a |
| **Healthy controls:**<br>**Contemporaneous**<br>**(Sheffield)** | 13 | 7/6<br>(54/46%) | 39 (32-46) | n/a | n/a |
| **Healthy controls: Pre-**<br>**Pandemic (Oxford) \*** | 12 | 6/4<br>(60/40%) | 25 (21-38) | n/a | n/a |
| **Healthy controls: Pre-**<br>**Pandemic (Liverpool)** | 48 | 35/13<br>(73/27%) | 26 (22-38.5) | n/a | n/a |
| **PCR Negative**<br>**Inpatients** | 9 | 4/5<br>(44/56%) | 74 (47-79) | n/a | n/a |
| **TOTAL Unexposed**<br>**controls** | 112 |  |  |  |  |
| **Highly Exposed - PCR**<br>**& Antibody negative** | 10 | 3/7<br>(30/70%) | 31 (25-34) | n/a | n/a |
| **P value (ANOVA)** |  | 0.002 | <0.0001 | <0.0001 | 0.03 |

\*missing demographic data for 2 samples

**Supplementary Table S2:** *Ex vivo* interferon-gamma ELISpot Responses by Group.

| Antigen stimulants | Spike |  |  | Structural & Accessory Proteins |  |  | Predicted Epitopes |  |  |
| --- | --- | --- | --- | --- | --- | --- | --- | --- | --- |
| Group | N | Median (IQR) SFC/10<sup>6</sup> | Res-ponder (%) | N | Median (IQR) SFC/10<sup>6</sup> | Res-ponder (%) | N | Median (IQR) SFC/10<sup>6</sup> | Res-ponder (%) |
| Asymptomatic SARS-CoV-2 infection | 22 | 0 (0-0) | 4/22 (18%) | 23 | 43 (0-148) | 12/23 (52%) | 2 | 28 (0-55) | 1/2 (50%) |
| Mild SARS-CoV-2 infection | 46 | 49 (0-207) | 27/46 (59%) | 73 | 108 (0-325) | 47/73 (64%) | 24 | 205 (0-381) | 21/24 (88%) |
| Severe / critical SARS-CoV-2 infection | 7 | 97 (0-919) | 3/7 (43%) | 7 | 113 (0-223) | 6/7 (86%) | 3 | 130 (0-1780) | 2/3 (67%) |
| Healthy seronegative controls | 22 | 0 (0-0) | 0/22 (0%) | 23 | 0 (0-0) | 1/23 (4%) | 5 | 0 (0-43) | 1/5 (20%) |
| Hospitalised PCR negative | 7 | 0 (0-0) | 0/7 (0%) | 6 | 0 (0-0) | 0/6 (0%) | 5 | 0 (0-0) | 0/5 (0%) |

**Supplementary Table S3:** *Ex vivo* interferon-gamma ELISpot Responses for Symptomatic versus Asymptomatic cases.

| Group | Symptomatic SARS-CoV-2 infection (mild, severe & critical) |  |  |  | Asymptomatic SARS-CoV-2 infection |  |  |  | P value (median ELISpot symptomatic v asymptomatic) |
| --- | --- | --- | --- | --- | --- | --- | --- | --- | --- |
|  | N | Median (IQR) SFC/10<sup>6</sup> | Res-ponder (%) | No. pools | N | Median (IQR) SFC/10<sup>6</sup> | Res-ponder (%) | No. pools response |  |
| Spike | 52 | 49 (0-216) | 30/52 (58%) | 1.3/12 (0-9) | 22 | 0 (0-0) | 4/22 (18%) | 0.4/12 (0-5) | 0.0051 |
| Structural & Accessory Proteins | 80 | 127 (0-335) | 53/80 (66%) | 1.2/7 (0-6) | 23 | 43 (0-148) | 12/23 (52%) | 1.0/7 (0-3) | 0.0378 |

**Supplementary Table S4:** *Ex vivo* interferon-gamma ELISpot responses amongst symptomatic Healthcare workers reporting fever versus no fever reported.

| Group | Fever reported by mild symptomatic SARS-CoV-2 infection |  |  |  | No fever reported by mild symptomatic SARS-CoV-2 infection |  |  |  | P value (median ELISpot fever v no fever) |
| --- | --- | --- | --- | --- | --- | --- | --- | --- | --- |
|  | N | Median (IQR) SFC/10<sup>6</sup> | Res-ponder (%) | No. pools | N | Median (IQR) SFC/10<sup>6</sup> | Res-ponder (%) | No. pools |  |
| Spike | 32 | 65 (0-226) | 22/32 (69%) | 1.5 (0-9) | 14 | 0 (0-110) | 5/14 (36%) | 0.7/12 (0-4) | 0.0269 |
| Structural & Accessory Proteins | 47 | 130 (0-355) | 31/47 (66%) | 1.4 (0-6) | 26 | 88 (0-146) | 16/26 (62%) | 0.6/7 (0-3) | 0.1008 |
| Predicted pools | 14 | 307 (0-399) | 13/14 (93%) | 2.2/4 (0-4) | 10 | 311 (45-639) | 8/10 (80%) | 1.7/4 (0-3) | 0.1549 |

**Supplementary Table S5.** Kruskal Wallis test with Dunn’s multiple comparison on magnitude of *ex vivo* interferon-gamma ELISpot responses.

|  | Dunn's multiple comparisons test | Summary of P value | Adjusted P Value |
| --- | --- | --- | --- |
| **SPIKE** | Control vs. Asymptomatic | ns | >0.9999 |
|  | Control vs. Mild HCW | **** | <0.0001 |
|  | Control vs. Severe/Critical | ns | 0.0777 |
|  | Control vs. Negative inpatient | ns | >0.9999 |
|  | Asymptomatic vs. Mild HCW | * | 0.0276 |
|  | Asymptomatic vs. Severe/Critical | ns | 0.6870 |
|  | Asymptomatic vs. Negative inpatient | ns | >0.9999 |
|  | Mild HCW vs. Severe/Critical | ns | >0.9999 |
|  | Mild HCW vs. Negative inpatient | * | 0.0394 |
|  | Severe/Critical vs. Negative inpatient | ns | 0.2678 |
| **STRUCTURAL & ACCESSORY** | Control vs. Asymptomatic | ns | 0.0984 |
|  | Control vs. Mild HCW | **** | <0.0001 |
|  | Control vs. Severe/Critical | *** | 0.0003 |
|  | Control vs. Negative inpatient | ns | >0.9999 |
|  | Asymptomatic vs. Mild HCW | ns | 0.8993 |
|  | Asymptomatic vs. Severe/Critical | ns | 0.1755 |
|  | Asymptomatic vs. Negative inpatient | ns | 0.7317 |
|  | Mild HCW vs. Severe/Critical | ns | >0.9999 |
|  | Mild HCW vs. Negative inpatient | * | 0.0386 |
|  | Severe/Critical vs. Negative inpatient | ** | 0.0090 |
| **PREDICTED** | Control vs. Asymptomatic | ns | >0.9999 |
|  | Control vs. Mild HCW | * | 0.0410 |
|  | Control vs. Severe/Critical | ns | >0.9999 |
|  | Control vs. Negative inpatient | ns | >0.9999 |
|  | Asymptomatic vs. Mild HCW | ns | 0.8620 |
|  | Asymptomatic vs. Severe/Critical | ns | >0.9999 |
|  | Asymptomatic vs. Negative inpatient | ns | >0.9999 |
|  | Mild HCW vs. Severe/Critical | ns | >0.9999 |
|  | Mild HCW vs. Negative inpatient | * | 0.0114 |
|  | Severe/Critical vs. Negative inpatient | ns | 0.8517 |
| **CEF POSITIVE CONTROL** | Control vs. Asymptomatic | ns | >0.9999 |
|  | Control vs. Mild HCW | ns | 0.7163 |
|  | Control vs. Severe/Critical | ns | 0.7528 |
|  | Control vs. Negative inpatient | ns | >0.9999 |
|  | Asymptomatic vs. Mild HCW | ns | 0.1164 |
|  | Asymptomatic vs. Severe/Critical | ns | 0.3007 |
|  | Asymptomatic vs. Negative inpatient | ns | 0.5417 |
|  | Mild HCW vs. Severe/Critical | ns | >0.9999 |
|  | Mild HCW vs. Negative inpatient | ns | >0.9999 |
|  | Severe/Critical vs. Negative inpatient | ns | >0.9999 |

**Supplementary Table S6a.** Kruskal Wallis test with Dunn’s multiple comparison on frequency of proliferative CD4+ T cells.

| Dunn's multiple comparisons test | Summary of P value | Adjusted P Value |
| --- | --- | --- |
| DMSO vs. S1 | **** | <0.0001 |
| DMSO vs. S2 | **** | <0.0001 |
| DMSO vs. M | **** | <0.0001 |
| DMSO vs. NP | **** | <0.0001 |
| DMSO vs. ORF 3 | **** | <0.0001 |
| DMSO vs. ORF 6 | ns | >0.9999 |
| DMSO vs. ORF 7 | ns | 0.0669 |
| DMSO vs. ORF 8 | ** | 0.0054 |
| S1 vs. S2 | ns | >0.9999 |
| S1 vs. M | ns | >0.9999 |
| S1 vs. NP | ns | >0.9999 |
| S1 vs. ORF 3 | *** | 0.001 |
| S1 vs. ORF 6 | **** | <0.0001 |
| S1 vs. ORF 7 | **** | <0.0001 |
| S1 vs. ORF 8 | **** | <0.0001 |
| S2 vs. M | ns | >0.9999 |
| S2 vs. NP | ns | >0.9999 |
| S2 vs. ORF 3 | ** | 0.0095 |
| S2 vs. ORF 6 | **** | <0.0001 |
| S2 vs. ORF 7 | **** | <0.0001 |
| S2 vs. ORF 8 | **** | <0.0001 |
| M vs. NP | ns | >0.9999 |
| M vs. ORF 3 | **** | <0.0001 |
| M vs. ORF 6 | **** | <0.0001 |
| M vs. ORF 7 | **** | <0.0001 |
| M vs. ORF 8 | **** | <0.0001 |
| NP vs. ORF 3 | *** | 0.0009 |
| NP vs. ORF 6 | **** | <0.0001 |
| NP vs. ORF 7 | **** | <0.0001 |
| NP vs. ORF 8 | **** | <0.0001 |
| ORF 3 vs. ORF 6 | *** | 0.0007 |
| ORF 3 vs. ORF 7 | ns | 0.1732 |
| ORF 3 vs. ORF 8 | ns | >0.9999 |
| ORF 6 vs. ORF 7 | ns | >0.9999 |
| ORF 6 vs. ORF 8 | ns | >0.9999 |
| ORF 7 vs. ORF 8 | ns | >0.9999 |

**Supplementary Table S6b.**
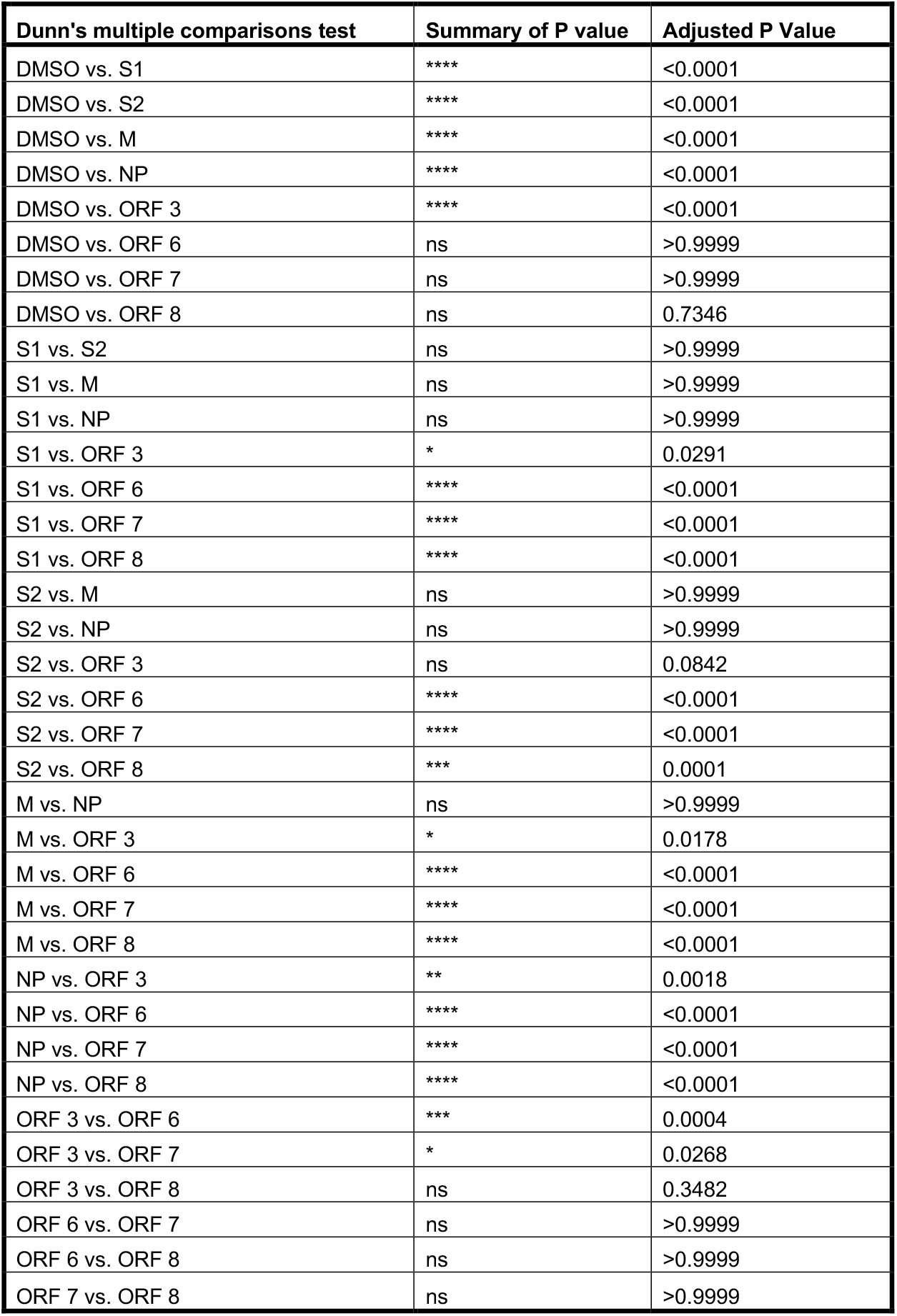
Kruskal Wallis test with Dunn’s multiple comparison on frequency of proliferative CD8+ T cells.

**Supplementary Table S7.**
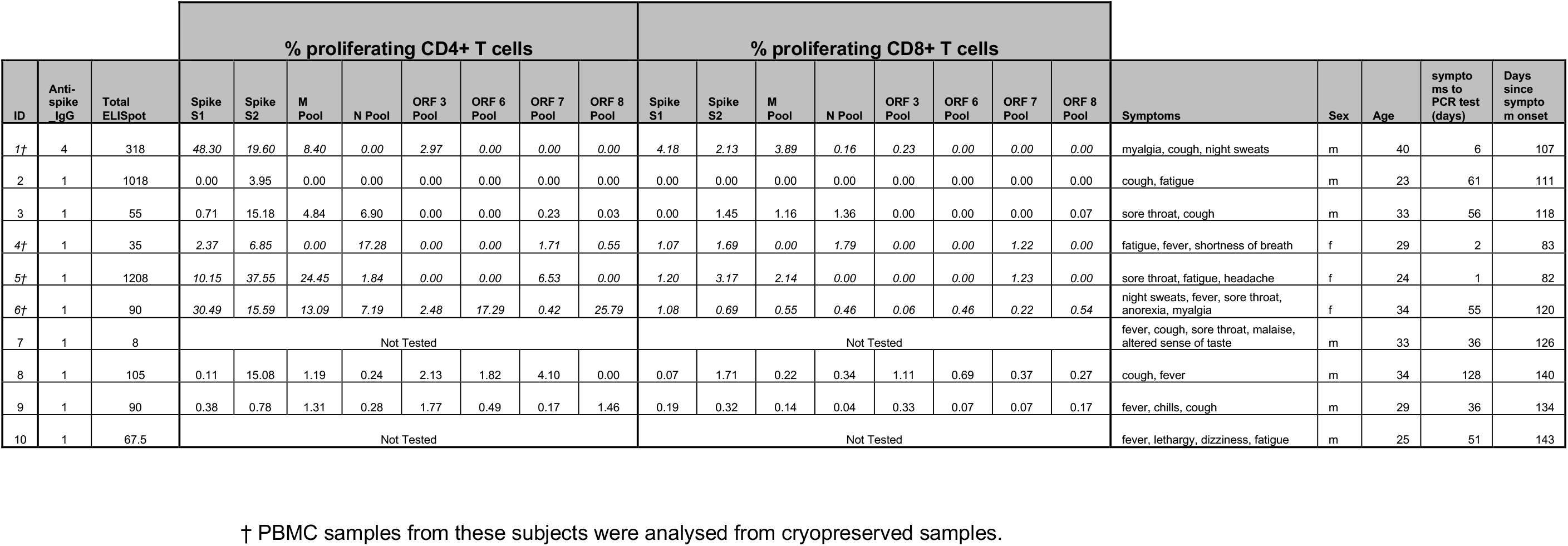
Clinical information and ELISpot and proliferation assay responses for highly exposed doctors used in this study.

|  |  |  | % proliferating CD4+ T cells |  |  |  |  |  |  |  | % proliferating CD8+ T cells |  |  |  |  |  |  |  | Symptoms | Sex | Age | symptoms to PCR test (days) | Days since symptom onset |
| --- | --- | --- | --- | --- | --- | --- | --- | --- | --- | --- | --- | --- | --- | --- | --- | --- | --- | --- | --- | --- | --- | --- | --- |
| ID | Anti-spike IgG | Total ELISpot | Spike S1 | Spike S2 | M Pool | N Pool | ORF 3 Pool | ORF 6 Pool | ORF 7 Pool | ORF 8 Pool | Spike S1 | Spike S2 | M Pool | N Pool | ORF 3 Pool | ORF 6 Pool | ORF 7 Pool | ORF 8 Pool |  |  |  |  |  |
| 1† | 4 | 318 | 48.30 | 19.60 | 8.40 | 0.00 | 2.97 | 0.00 | 0.00 | 0.00 | 4.18 | 2.13 | 3.89 | 0.16 | 0.23 | 0.00 | 0.00 | 0.00 | myalgia, cough, night sweats | m | 40 | 6 | 107 |
| 2 | 1 | 1018 | 0.00 | 3.95 | 0.00 | 0.00 | 0.00 | 0.00 | 0.00 | 0.00 | 0.00 | 0.00 | 0.00 | 0.00 | 0.00 | 0.00 | 0.00 | 0.00 | cough, fatigue | m | 23 | 61 | 111 |
| 3 | 1 | 55 | 0.71 | 15.18 | 4.84 | 6.90 | 0.00 | 0.00 | 0.23 | 0.03 | 0.00 | 1.45 | 1.16 | 1.36 | 0.00 | 0.00 | 0.00 | 0.07 | sore throat, cough | m | 33 | 56 | 118 |
| 4† | 1 | 35 | 2.37 | 6.85 | 0.00 | 17.28 | 0.00 | 0.00 | 1.71 | 0.55 | 1.07 | 1.69 | 0.00 | 1.79 | 0.00 | 0.00 | 1.22 | 0.00 | fatigue, fever, shortness of breath | f | 29 | 2 | 83 |
| 5† | 1 | 1208 | 10.15 | 37.55 | 24.45 | 1.84 | 0.00 | 0.00 | 6.53 | 0.00 | 1.20 | 3.17 | 2.14 | 0.00 | 0.00 | 0.00 | 1.23 | 0.00 | sore throat, fatigue, headache | f | 24 | 1 | 82 |
| 6† | 1 | 90 | 30.49 | 15.59 | 13.09 | 7.19 | 2.48 | 17.29 | 0.42 | 25.79 | 1.08 | 0.69 | 0.55 | 0.46 | 0.06 | 0.46 | 0.22 | 0.54 | night sweats, fever, sore throat, anorexia, myalgia | f | 34 | 55 | 120 |
| 7 | 1 | 8 | Not Tested |  |  |  |  |  |  |  | Not Tested |  |  |  |  |  |  |  | fever, cough, sore throat, malaise, altered sense of taste | m | 33 | 36 | 126 |
| 8 | 1 | 105 | 0.11 | 15.08 | 1.19 | 0.24 | 2.13 | 1.82 | 4.10 | 0.00 | 0.07 | 1.71 | 0.22 | 0.34 | 1.11 | 0.69 | 0.37 | 0.27 | cough, fever | m | 34 | 128 | 140 |
| 9 | 1 | 90 | 0.38 | 0.78 | 1.31 | 0.28 | 1.77 | 0.49 | 0.17 | 1.46 | 0.19 | 0.32 | 0.14 | 0.04 | 0.33 | 0.07 | 0.07 | 0.17 | fever, chills, cough | m | 29 | 36 | 134 |
| 10 | 1 | 67.5 | Not Tested |  |  |  |  |  |  |  | Not Tested |  |  |  |  |  |  |  | fever, lethargy, dizziness, fatigue | m | 25 | 51 | 143 |

† PBMC samples from these subjects were analysed from cryopreserved samples.

## Figure legends

**Supplementary Figure 1. Detailed SARS-CoV-2 specific Immune Response**

**a)** Neutralising antibody responses in a pseudoparticle assay in seronegative controls, asymptomatic and mildly symptomatic healthcare workers (HCWs) with PCR-confirmed SARS-CoV-2 infection, hospitalised patients with severe or critical PCR-confirmed SARS-CoV-2 infection, and PCR-negative inpatient controls. *Ex vivo* IFN-γ ELISpot showing the magnitude and breadth of effector T cell responses to **b)** SARS-CoV-2 spike peptide pools and M, N, and accessory proteins ORF 3, ORF6, ORF7 and ORF8 and **c)** *in silico* predicted pools^10^ in HCWs convalescent with mildly symptomatic SARS-CoV-2 infection. X axis shows number of days from onset of symptoms (not to scale), with blank columns representing zero response in the subject tested at that time-point. **d)** Correlation between summed *ex vivo* IFN-γ ELISpot responses to spike protein and to structural and accessory proteins. Correlation between days from symptom onset and summed *ex vivo* IFN-γ ELISpot responses to **e)** spike protein and **f)** structural and accessory proteins. *Ex vivo* IFN-γ ELISpot responses to summed SARS-CoV-2 peptide pools spanning spike, accessory and structural proteins (E, M, N, ORF 3, ORF6, ORF7 and ORF8), *in silico* predicted pools^10^ and the CEF T cell control panel by **g)** age band in years and by **h)** sex. SFC/10^6^ PBMC = spot forming cells per million peripheral blood mononuclear cells, with background subtracted. Correlation was performed via Spearman’s rank correlation coefficient and comparison of three groups for age by Kruskal-Wallis one-way ANOVA and two groups for sex by Mann-Whitney U test. ns = not significant, * = <0.05, ** = <0.01, *** = < 0.001 and **** = <0.0001.

**Supplementary Figure 2. Gating strategy and representative plots for CTV assay. a)** gating strategy for identification of proliferating CD4+ and CD8+ T cells. **b)** Representative plots for gating peptide pool specific proliferative responses. **c)** Representative plots for validation of specificity of proliferation assay using HCV seronegative samples **d)** quantification of HCV and FEC-T specific protein response in CD4+ and **e)** CD8+ T cells. PHA is used as positive control. All data have been background subtracted. Data shows media with interquartile range

**Supplementary Figure 3. Comparative analysis of the magnitude of proliferative responses** in **a)** CD4+ and CD8+ T cells. **b)** mild but symptomatic and asymptomatic convalescent HCWs within the CD4+ T cell population and **c)** mild but symptomatic and asymptomatic convalescent HCWs within the CD8+ T cell population

**Supplemental Figure 4. Representative ICS plots**.

PBMC were stimulated with 2ug/mL of the indicated peptide pool or DMSO control. Representative plots for gated CD8+ T cells are shown in A and for gated CD4+T cells in B.

**Supplemental Fig 5. ICS responses in CD4+ and CD8+ T cells for Spike Pools**.

ICS was performed as in Figure 4 on n=26 individuals PCR+ for SARS-CoV-2. Expression levels of IFN-γ, IL-2 and TNF-α in CD4+ and CD8+ T cells are shown for the peptide pools M in (**a**), NP in (**d**), S1 in (**g**), and S2 in (**j**). Bars represent median +/- IQR. Statistics were performed using Wilcoxon matched-pairs signed rank test between each cytokine in CD4+ vs CD8+ T cells. Polyfunctionality was then assessed as in Figure 4. Polyfunctionality for M pools is shown for CD4+ T cells in (**b**) and CD8+ T cells in (**c**). **e)** and **f)** show polyfunctionality with NP pools in CD4+ and CD8+ T cells respectively. Polyfunctional responses with S1 pools are shown for CD4+ (**h**) and CD8+ T cells in (**i**). Polyfunctionality analysis was also performed with S2 pools and are shown for CD4+ (**k**) and CD8+ T cells (**l**).

**Supplementary Figure 6. Comparative analysis of the magnitude of proliferative responses** in SARS-COV-2 seronegative controls from 2020 analysed from fresh PBMCs and cryopreserved pre-pandemic seronegative controls in **a)** CD4+ and **b)** CD8+ T cells.

